# Risk factors mediating the effect of body-mass index and waist-to-hip ratio on cardiovascular outcomes: Mendelian randomization analysis

**DOI:** 10.1101/2020.07.15.20154096

**Authors:** Dipender Gill, Verena Zuber, Jesse Dawson, Jonathan Pearson-Stuttard, Alice R. Carter, Eleanor Sanderson, Ville Karhunen, Michael G. Levin, Robyn E. Wootton, VA Million Veteran Program, Derek Klarin, Philip S. Tsao, Konstantinos K. Tsilidis, Scott M. Damrauer, Stephen Burgess, Paul Elliott

## Abstract

**Background:** Higher body-mass index (BMI) and waist-to-hip ratio (WHR) increase the risk of cardiovascular disease, but the extent to which this is mediated by blood pressure, diabetes, lipid traits and smoking is not fully understood.

**Methods:** Using consortia and UK Biobank genetic association summary data from 140,595 to 898,130 participants predominantly of European ancestry, MR mediation analysis was performed to investigate the degree to which genetically predicted systolic blood pressure (SBP), diabetes, lipid traits and smoking mediated an effect of genetically predicted BMI and WHR on risk of coronary artery disease (CAD), peripheral artery disease (PAD) and stroke.

**Results:** The 49% (95% confidence interval [CI] 39%-60%) increased risk of CAD conferred per 1-standard deviation increase in genetically predicted BMI attenuated to 34% (95% CI 24%-45%) after adjusting for genetically predicted SBP, to 27% (95% CI 17%-37%) after adjusting for genetically predicted diabetes, to 47% (95% CI 36%-59%) after adjusting for genetically predicted lipids, and to 46% (95% CI 34%-58%) after adjusting for genetically predicted smoking. Adjusting for all the mediators together, the increased risk attenuated to 14% (95% CI 4%-26%). A similar pattern of attenuation was observed when considering genetically predicted WHR as the exposure, and PAD or stroke as the outcomes.

**Conclusions:** Measures to reduce obesity will lower risk of cardiovascular disease primarily by impacting on downstream metabolic risk factors, particularly diabetes and hypertension. Reduction of obesity prevalence alongside control and management of its mediators is likely to be most effective for minimizing the burden of obesity.

## Background

Cardiovascular disease (CVD) is the leading cause of death and disability worldwide(1). Obesity can contribute towards CVD risk through effects on hyperglycaemia, hypertension, dyslipidaemia, and smoking behaviour(2-5). The global prevalence of obesity has more than tripled in the last 40 years, with an even greater rise in incidence amongst children(6). It is estimated that by 2030, approximately half of the US population will be obese(7). There are treatments available to effectively manage the downstream mediators through which obesity causes CVD(8-11). Understanding of such pathways is therefore paramount to reducing cardiovascular risk.

Obesity can be measured by various means, and is often defined as a body-mass index (BMI) of greater than 30kg/m^2^ (12). However, BMI is a not a direct measure of adiposity, and is also correlated with fat-free mass(12). Assessment of obesity using waist-to-hip ratio (WHR) is less subject to influence from height and muscle mass, and is associated with cardiovascular risk in individuals with a normal BMI(13, 14). Both BMI and WHR are easy to measure clinically, without any requirement for radiological investigation. Conventional observational studies have shown that the relationship between obesity measures such as BMI and WHR with CVD is attenuated when adjustment is made for cardiometabolic risk factors such as blood pressure, lipid traits or measures of glycaemia(15). This has allowed for estimation of the proportion of the effect of obesity that is mediated through these intermediates(15). However, such analysis is vulnerable to bias from confounding and measurement error, both of which can result in underestimation of the proportion of effect mediated(16, 17). The Mendelian randomization (MR) approach uses genetic variants as instruments for studying the effect of an exposure on an outcome, and has now been extended to perform mediation analyses(16, 18). The use of randomly allocated genetic variants in this paradigm means that the estimates generated are less vulnerable to confounding from environmental factors, with consideration of their lifelong effects reducing bias from measurement error(16).

The increasing availability of large-scale genome-wide association study (GWAS) data has greatly facilitated MR analyses considering cardiovascular risk factors and outcomes. In this study, we aimed to use such data within the MR framework to investigate the role of blood pressure, diabetes, fasting glucose, lipid traits and smoking in mediating the effect of BMI and WHR respectively on coronary artery disease (CAD), peripheral arterial disease (PAD) and stroke risk.

## Methods

### Ethical approval, data availability and reporting

The data used in this work are publicly available and the studies from which they were obtained are cited. All these studies obtained relevant participant consent and ethical approval. The results from the analyses performed in this work are presented in the main manuscript or its supplementary files. This paper has been reported based on recommendations by the STROBE-MR Guidelines (Research Checklist)(19). The study protocol and details were not pre-registered.

### Data sources

Genetic association estimates for BMI and WHR were obtained from the GIANT Consortium GWAS meta-analysis of 806 834 and 697 734 European-ancestry individuals respectively(20). Genetic association estimates for SBP were obtained from a GWAS of 318 417 White British individuals in the UK Biobank, with correction made for any self-reported anti-hypertensive medication use by adding 10mmHg to the mean SBP measured from two automated recordings that were taken two minutes apart at baseline assessment(21). Genetic association estimates for lifetime smoking (referred to hereon as smoking) were obtained from a GWAS of 462 690 European-ancestry individuals in the UK Biobank(22). A lifetime measure of smoking was created based on self-reported age at initiation, age at cessation and cigarettes smoked per day(22). Genetic association estimates for liability to diabetes came from the DIAGRAM Consortium GWAS meta-analysis of 74 124 cases and 824 006 controls, all of European ancestry(23). Genetic association estimates for plasma fasting glucose were obtained by using PLINK software to carry out a meta-analysis of MAGIC Consortium GWAS summary data from separate analyses of 67 506 men and 73 089 women who were not diabetic(24, 25). Genetic association estimates for fasting serum low-density lipoprotein cholesterol (LDL-C), high-density lipoprotein cholesterol (HDL-C) and triglycerides were obtained from the Global Lipids Genetic Consortium GWAS of 188,577 European-ancestry individuals(26). Genetic association estimates for CAD were obtained from the CARDIoGRAMplusC4D Consortium 1000G multi-ethnic GWAS (77% European-ancestry) of 60 801 cases and 123 504 controls(27). Genetic association estimates for PAD were obtained from the Million Veterans Program multi-ethnic (72% European-ancestry) GWAS of 31 307 cases and 211 753 controls(28). Genetic association estimates for stroke were obtained from the MEGASTROKE multi-ethnic (86% European-ancestry) GWAS of 67 162 cases (of any stroke) and 454 450 controls(29). Population characteristics and specific trait definitions relating to all these summary genetic association estimates are available in their original publications. For the analyses performed in this current work, genetic variants from different studies were aligned by their effect alleles and no exclusions were made for palindromic variants. Only variants for which genetic association estimates were available for all the traits being investigated in any given analysis were considered, and proxies were not used.

### Instrument selection

To estimate the total effect of BMI and WHR respectively on the considered cardiovascular outcomes, instruments were selected as single-nucleotide polymorphisms (SNPs) that associated with BMI or WHR at genome-wide significance (*P*<5×10^−8^) and were in pair-wise linkage disequilibrium (LD) *r*^2^<0.001. To select instruments for mediation analysis, all SNPs related to the considered exposure (BMI or WHR) or mediators at genome-wide significance were pooled and clumped to pairwise LD *r*^2^<0.001 based on the lowest *P*-value for association with any trait. All clumping was performed using the TwoSampleMR package in R(30).

### Total effects

Random-effects inverse-variance weighted (IVW) MR was used as the main analysis for estimating the total effects of genetically predicted BMI and genetically predicted WHR respectively on each of the considered CVD outcomes(31). Contamination-mixture method and weighted median MR were used in sensitivity analyses to explore the robustness of the findings to potential pleiotropic effects of the variants(32, 33). The contamination-mixture model makes the assumption that MR estimates from valid instruments follow a normal distribution that centres on the true causal effect estimate, while those calculated from invalid instrument variants follow a normal distribution centred on the null(33). This allows for a likelihood function to be specified and maximized when allocating each variant to one of the two mixture distributions(33). The weighted median approach orders the MR estimates from individual variants by their magnitude weighted for their precision and selects the median as the overall MR estimate, calculating standard error by bootstrapping(32). The MendelianRandomization package in R was used for performing the IVW, contamination-mixture and weighted median MR analyses(34).

### Mediation analysis

To estimate the direct effect of genetically predicted BMI and genetically predicted WHR on each of the three considered CVD outcomes that was not being mediated by the investigated intermediary risk factors, summary data multivariable MR was performed(35-37). Specifically, the variant-outcome genetic association estimates were regressed on the variant-exposure and variant-mediator estimates, weighted for the precision of the variant-outcome association, and with the intercept fixed to zero(37). Using this approach, adjustment was made for genetically predicted SBP, diabetes, smoking and lipid traits (LDL-C, HDL-C and triglycerides together) in turn, and finally including all mediators together in a joint model. In a sensitivity analysis, genetically predicted diabetes was excluded from this joint model to remove any bias that might be introduced because of its binary nature(38). For analyses considering genetically predicted fasting glucose in non-diabetics instead of genetically predicted diabetes, the corresponding genetic association data were substituted.

Multivariable MR mediation analysis was performed to estimate the proportion of the effect of BMI and WHR respectively on CAD, PAD and stroke that was mediated through each of the considered risk factors, and also all of them together(16). Specifically, the direct effect of genetically predicted BMI and genetically predicted WHR respectively was divided by their total effect and subtracted from 1, with standard errors estimated using the propagation of error method(16, 18).

### Independent effects of genetically predicted BMI and WHR

The direct effects of genetically predicted BMI and genetically predicted WHR on the considered CVD outcomes that are not mediated through each other were measured by including only these two traits together as exposures in the summary data multivariable MR model described above.

## Results

### Total effects

Considering total effects, there was consistent evidence across the IVW, contamination-mixture and weighted median MR methods that both higher genetically predicted BMI and higher genetically predicted WHR increased CAD, PAD and stroke risk (Figure 1). In the main IVW MR analysis, the odds ratio (OR) per 1-standard deviation (SD) increase in genetically predicted BMI for CAD risk was 1.49 (95% confidence interval [CI] 1.39-1.60), for PAD risk was 1.70 (95% CI 1.58-1.82), and for stroke risk was 1.22 (95% CI 1.15-1.29). For a 1-SD increase in genetically predicted WHR, this was 1.54 (95% CI 1.38-1.71) for CAD risk, 1.55 (95% CI 1.40-1.71) for PAD risk, and 1.30 (95% CI 1.21-1.40) for stroke risk.

**Figure 1.**
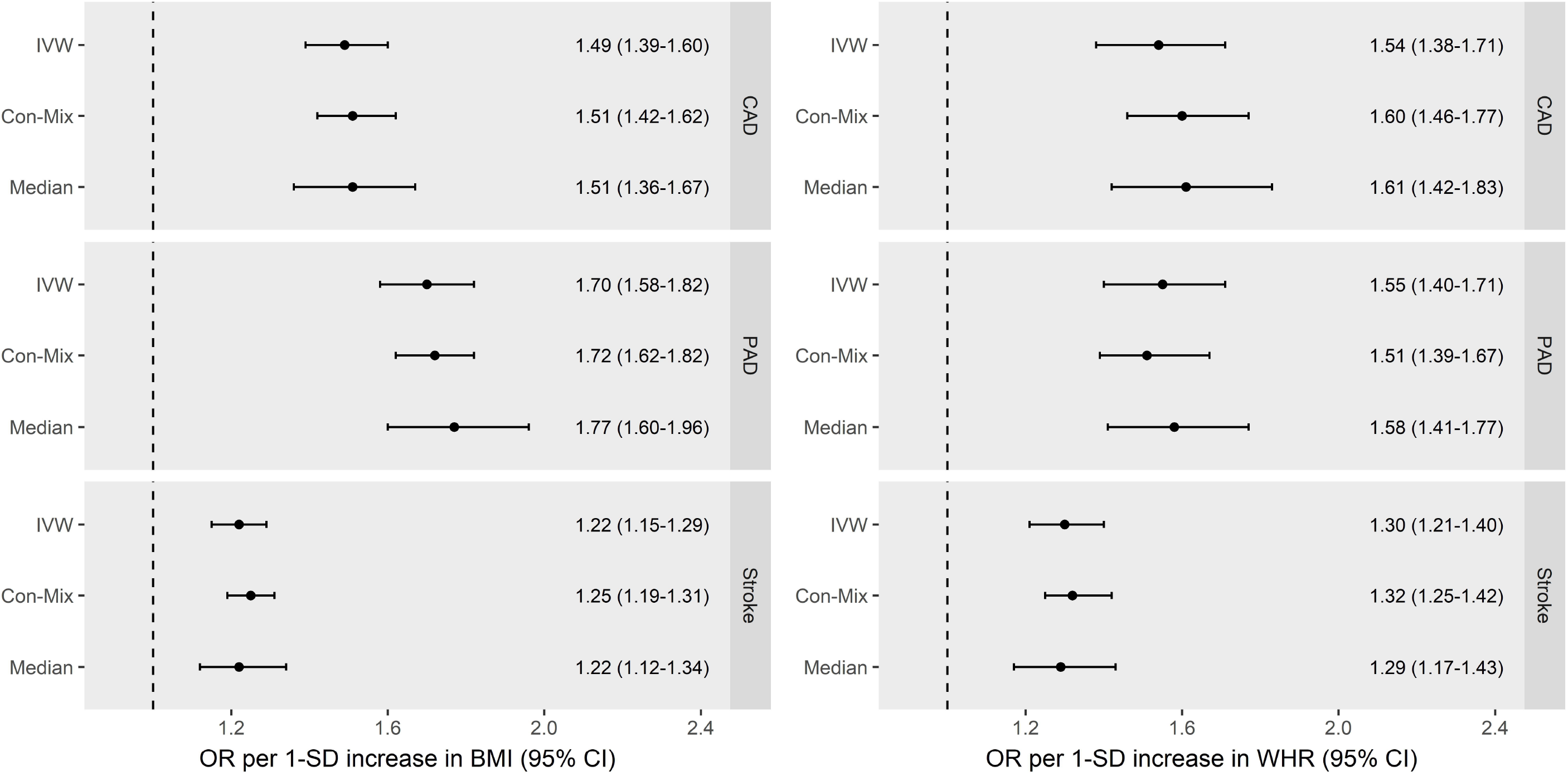
Total effects of genetically predicted body mass index (BMI) and genetically predicted waist-to-hip ratio (WHR) on coronary artery disease (CAD), peripheral artery disease (PAD) and stroke. Inverse-variance weighted (IVW), contamination-mixture method (Con-Mix) and weighted median (Median) represent distinct Mendelian randomization approaches that differ in their requisite assumptions. CI: confidence interval; IVW: inverse-variance weighted; OR: odds ratio; SD: standard deviation.

### Mediation analysis

There was attenuation in the effect of genetically predicted BMI and genetically predicted WHR on the three CVD outcomes after adjusting for genetically predicted SBP, diabetes, lipid traits (LDL-C, HDL-C and triglycerides together) and smoking, either separately or in the same joint model (Figure 2). The 49% (95% CI 39%-60%) increased risk of CAD conferred per 1-SD increase in genetically predicted BMI attenuated to 34% (95% CI 24%-45%) after adjusting for genetically predicted SBP, to 27% (95% CI 17%-37%) after adjusting for genetically predicted diabetes, to 47% (95% CI 36%-59%) after adjusting for genetically predicted lipids, and to 46% (95% CI 34%-58%) after adjusting for genetically predicted smoking. Adjusting for all the mediators together in the same model, the attenuation was to 14% (95% CI 4% to 26%).

**Figure 2.**
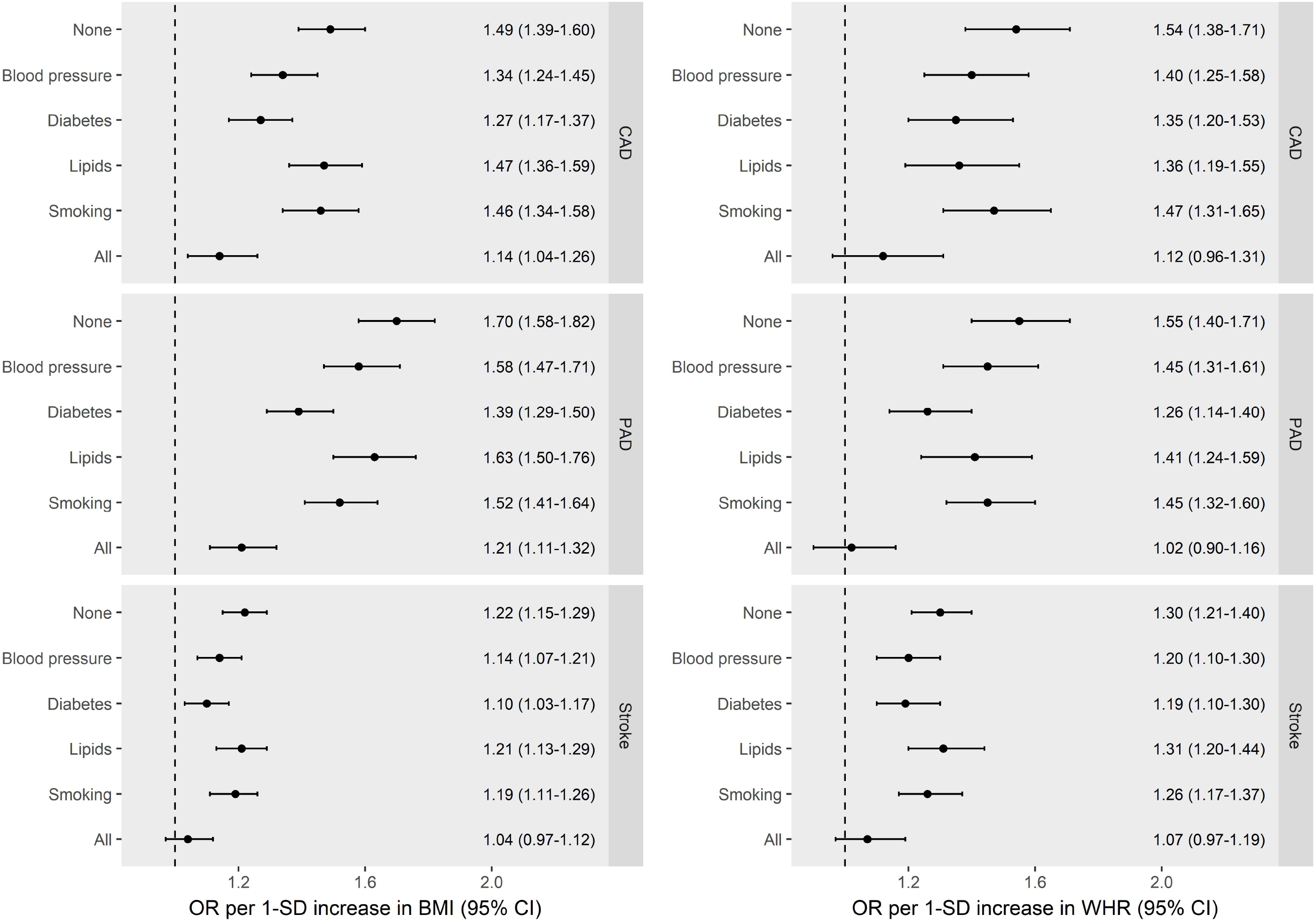
Direct effects of genetically predicted body mass index (BMI) and genetically predicted waist-to-hip ratio (WHR) on coronary artery disease (CAD), peripheral artery disease (PAD) and stroke, estimated after adjusting for genetic liability to mediators separately and together in the same model. The y-axis details the genetically predicted mediator(s) for which adjusted was made. Blood pressure relates to systolic blood pressure. Lipids relates to serum low-density lipoprotein cholesterol, high-density lipoprotein cholesterol and triglycerides considered together in one model. CI: confidence interval; OR: odds ratio; SD: standard deviation.

The percentage attenuation in the total effects of genetically predicted BMI and WHR respectively on the three CVD outcomes after adjusting for the mediators is depicted in Supplementary Figure 1. For the effect of genetically predicted BMI on CAD risk, 27% (95% CI 3%-50%) was mediated by genetically predicted SBP, 41% (95% 18%-63%) was mediated by genetically predicted diabetes, 3% (-23%-29%) was mediated by genetically predicted lipids, and 6% (95% CI -20%-32%) was mediated by genetically predicted smoking. All the mediators together accounted for 66% (95% CI 42%-91%) of the total effect of genetically predicted BMI on CAD risk.

A joint model including all considered mediators except genetically predicted diabetes was also constructed (Supplementary Figure 2). Adjusting together for all the mediators except genetically predicted diabetes, the effect of genetically predicted BMI on CAD risk attenuated from 49% (95% CI 39%-60%) to 27% (95% CI 16% to 40%).

There was little change in the association of either genetically predicted BMI or genetically predicted WHR with risk of the three CVD outcomes after adjusting for genetically predicted fasting glucose in non-diabetic individuals (Figure 3).

**Figure 3.**
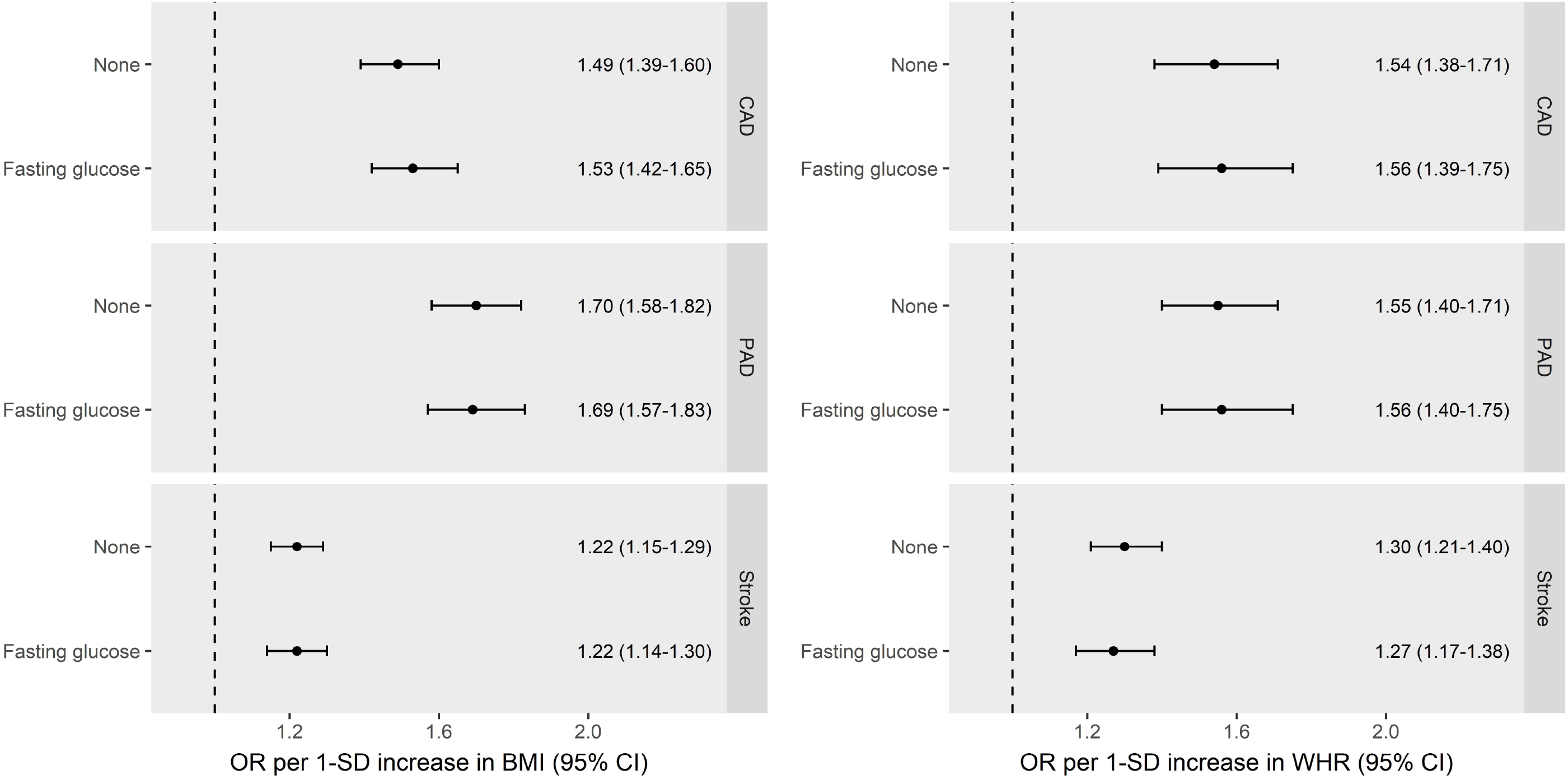
Direct effects of body mass index (BMI) and waist-to-hip ratio (WHR) on coronary artery disease (CAD), peripheral artery disease (PAD) and stroke, estimated after no adjustment and after adjustment for genetically predicted fasting glucose in non-diabetics. CI: confidence interval; OR: odds ratio; SD: standard deviation.

### Independent effects of genetically predicted BMI and WHR

Both genetically predicted BMI and genetically predicted WHR had direct effects on CAD, PAD and stroke after mutual adjustment (Figure 4). The increased CAD risk attributed to a 1-SD higher genetically predicted BMI attenuated from 49% (95% CI 39%-60%) to 32% (95% CI 20%-45%) after adjusting for genetically predicted WHR, and the increased CAD risk attributed to a 1-SD higher genetically predicted WHR attenuated from 54% (95% CI 38%-71%) to 33% (95% CI 18%-50%) after adjusting for genetically predicted BMI.

**Figure 4.**
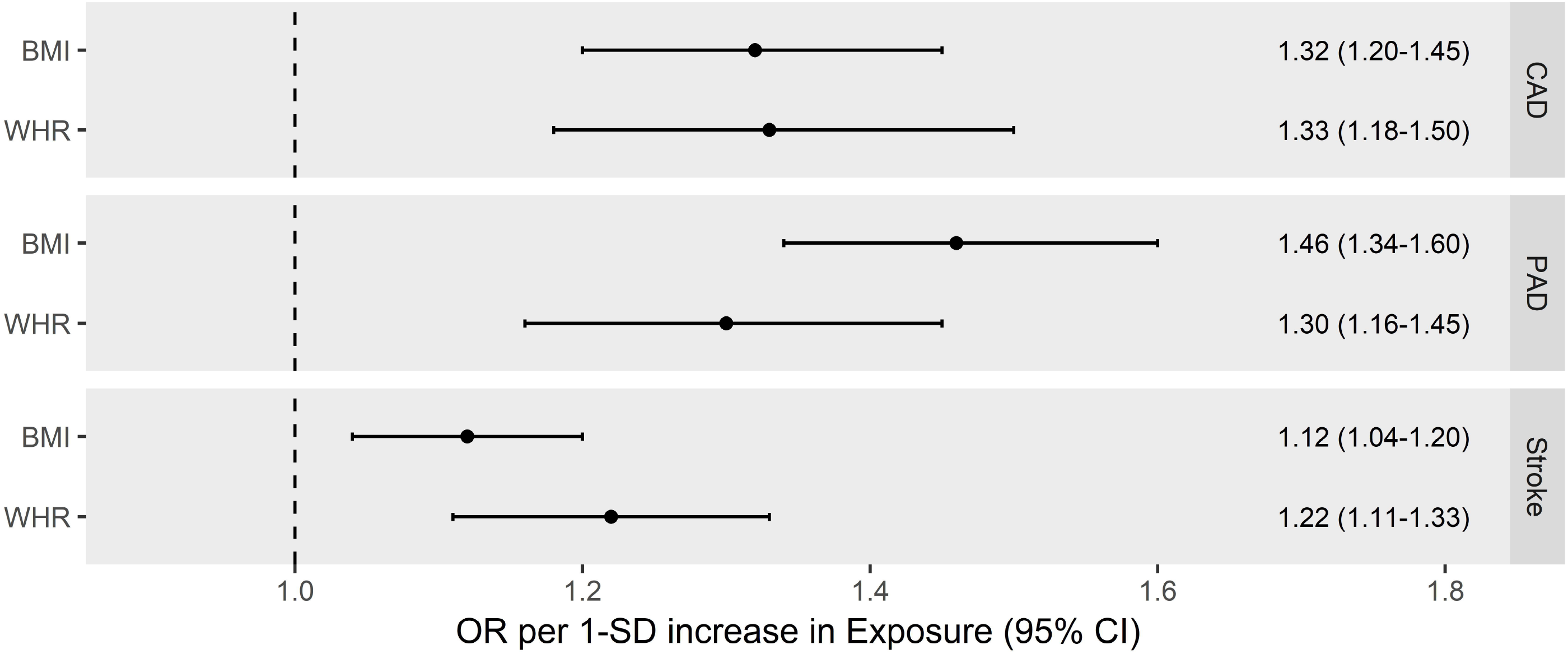
Direct effects of genetically predicted body mass index (BMI) and genetically predicted waist-to-hip ratio (WHR) on coronary artery disease (CAD), peripheral artery disease (PAD) and stroke, estimated after adjusting for each other. CI: confidence interval; OR: odds ratio; SD: standard deviation.

## Discussion

This study uses large-scale genetic association data within the MR paradigm to investigate the role of SBP, diabetes, lipid traits and smoking in mediating the effect of BMI and WHR on CAD, PAD and stroke risk. The results support the idea that the majority of the effects of obesity on CVD are mediated through these risk factors, with diabetes and blood pressure being the most notable and accounting for approximately one-third and one-quarter of the effect respectively. In contrast, the analysis of genetically predicted fasting glucose in non-diabetic individuals did not provide any evidence to support its role in mediating the effect of obesity on CVD risk.

The sum of the estimated mediating effects of the various risk factors considered individually was comparable to their total mediating effect estimated when considering them all together in the same model, consistent with them acting through distinct mechanisms. Including genetically predicted BMI and genetically predicted WHR in the same model produced evidence consistent with these traits having direct effects on CVD risk independently of each other. It follows that rather than analysing BMI or WHR alone, they should be considered together as they capture different aspects of adiposity.

Our findings have important clinical and public health implications. Behavioural interventions to reduce obesity can have inadequate long term effects(39), pharmacological treatments may be limited by unfavourable adverse effect profiles(40), and surgical procedures are often reserved for only severe cases(41). This work supports the concept that the majority of the cardiovascular consequences of obesity may be managed by effectively controlling its downstream mediators, most notably diabetes and raised blood pressure, for which effective pharmacological interventions are available. This has relevance for the more than 640 million individuals worldwide currently living with obesity(42), and the many more forecasted to become obese in coming years(43). Such holistic consideration of obesity together with its mediators could contribute to a shift from the single-disease focus of health systems towards prioritizing multi-morbidity and promoting individual and societal wellness(44). Our analyses were also suggestive of some possible residual effect of BMI and WHR on CVD risk even after adjusting for all the considered mediating risk factors, consistent with metabolically healthy obesity still conferring increased CVD risk(45). Taken together, these results suggest that unless the growing obesity epidemic is effectively tackled, we risk undoing the large reductions in CVD mortality achieved over past decades(1). Population-based approaches that decrease obesity by addressing key upstream drivers such as poor diet and physical inactivity have substantial potential for impact and are also effective for reducing health inequalities(46, 47).

The results of our current study can be contrasted to those from a large-scale observational analysis of 1.8 million people across 97 studies(15, 48). This previous work estimated that 46% (95% CI 42%-50%) of the excess risk conferred by raised BMI on CAD and 76% (95% CI 65%-91%) on stroke were mediated by effects on blood pressure, glucose levels and lipid traits, with blood pressure being the most important and mediation for stroke being greatest(15). However, the approach and data used in our current study offers a number of possible improvements. Our work includes a greater repertoire of risk factors and CVD outcomes than have been considered together previously(15, 49), in particular drawing on recently available GWAS summary data to study smoking and PAD(22, 28). MR analysis uses randomly allocated genetic variants that represent lifelong cumulative liability to the traits for which they serve as instruments and can therefore help overcome the confounding and measurement error that typically bias conventional observational studies(16). Consistent with this, our MR results indicate that these risk factors mediate a greater proportion of the effect of obesity than suggested by previous conventional observational analyses^(15)^.

Also of relevance here, we considered genetic liability to diabetes and genetically predicted fasting glucose in non-diabetic individuals as separate risk factors. Our findings support the concept that obesity traits confer an increased risk of CVD specifically through liability to diabetes, rather than variation in fasting glucose levels within the normal physiological range. This is important because fasting glucose may have a non-linear association with CVD risk(50), only having detrimental effects beyond a certain point(51).

Our current study also has limitations. The genetic association data used in this work are drawn from predominantly European populations, and should therefore be interpreted with caution when extrapolating to other ethnic groups. Diabetes is a binary outcome, and as such our consideration of genetically predicted diabetes could introduce bias into the mediation analysis because not all individuals possessing such genetic liability develop diabetes-related traits(38). SBP was used as a proxy for studying the effects of blood pressure more generally. Given the high degree of phenotypic and genetic correlation between blood pressure traits(52), this would seem unlikely to affect the conclusions drawn. A theoretical weakness of the MR approach relates to bias from pleiotropic effects of the genetic variants incorporated as instruments for the traits under study, whereby they may directly affect the outcome through pathways independent of the exposure or mediators being considered. Although such bias cannot be entirely excluded, it is reassuring that we obtained similar MR estimates for the total effect of BMI and WHR respectively on the three CVD outcomes when performing the IVW, contamination-mixture and weighted median MR methods that each make different assumptions concerning the presence of pleiotropic variants(53). Finally, there is currently no available method for assessing instrument strength within the two-sample multivariable MR setting, and we could therefore not assess potential vulnerability to weak instrument bias(35).

In conclusion, this work using the MR framework suggests that the majority of the effects of obesity on CVD risk are mediated through metabolic risk factors, most notably diabetes and blood pressure. Comprehensive public health measures that target the reduction of obesity prevalence alongside control and management of its downstream mediators are likely to be most effective for minimizing the burden of obesity on individuals and health systems alike.

## Data Availability

The results from the analyses performed in this work are presented in the main manuscript or its supplementary files.

## Author contributions

DG, JD, KKT, SMD and SB designed the project. DK, PST, SMD and VA-MVP provided data. DG and VZ analysed the data. DG, JD and JP-S drafted the manuscript. All authors interpreted the results and critically revised the manuscript. All authors approved the submitted article. All authors are accountable for the integrity of the research.

## Acknowledgements

The authors acknowledge the contributors of the data used in this work: CARDIoGRAMplusC4D, DIAGRAM, GIANT, Global Lipids Genetics Consortium, MAGIC, MEGASTROKE, Million Veterans Program and UK Biobank.

